# Computed Tomography Orodental Sub-volume Imaging Annotation Dataset of Head and Neck Cancer Radiation Therapy Patients

**DOI:** 10.1101/2025.09.09.25333678

**Authors:** Zaphanlene Kaffey, Austin H. Castelo, Renjie He, Lisanne V. van Dijk, Dong Joo Rhee, Congjun Wang, He C. Wang, Kareem A. Wahid, Sonali Joshi, Parshan Gerafian, Natalie A. West, Sarah Mirbahaeddin, Jaqueline Curiel, Samrina Acharya, Amal Shekha, Praise Oderinde, Alaa M.S. Ali, Andrew Hope, Erin Watson, Ruth Aponte-Wesson, Steven J. Frank, Carly E.A. Barbon, Kristy K. Brock, Mark S. Chambers, Muhammad Walji, Katherine A. Hutcheson, Stephen Y. Lai, Clifton D. Fuller, Mohamed A. Naser, Amy C. Moreno, Laia Humbert-Vidan, the OPC-SURVIVOR Program, the MD Anderson Head and Neck Cancer Symptom Working Group

## Abstract

Accurate delineation of orodental structures on computed tomography (CT) is critical for image-guided assessments of radiation-associated bone injury. This dataset comprises curated CT imaging and expert-defined segmentation masks for 60 patients with head and neck cancer treated with radiotherapy (RT), including delineations of mandibular and maxillary sub-volumes and individual teeth. Segmentation guidelines were informed by anatomical differences across sub-regions and aligned with the ClinRad osteoradionecrosis (ORN) staging system. The dataset includes converted NIfTI files of simulation CT images, RT dose distributions, and delineated structures. All segmentations were performed manually using a standardized protocol in a commercial treatment planning system and converted to research-ready formats using open-source tools. This dataset may facilitate the development and validation of automated segmentation tools, dose mapping applications, and image-based ORN detection pipelines in head and neck cancer survivors.

## Background & Summary

Radiotherapy (RT) plays a central role in the management of head and neck cancers (HNC), but unintended dose to adjacent bony structures such as the mandible and maxilla can result in long-term complications, including osteoradionecrosis (ORN). Clinical decision-making in this setting requires accurate imaging-based assessment of the jaws, including pre-existing dental conditions, post-treatment anatomical changes, and localized radiation dose exposure.

Existing automated segmentation methods for mandible^1–3^ and teeth^3–8^ are mostly focused on cone-beam computed tomography (CBCT) images and disregard the bone composition and radiobiological heterogeneity of the mandible. To address this gap, a mandible and maxilla sub-volumes and individual teeth deep-learning auto-segmentation model was recently developed using CT images to support decision-making in radiation oncology for head and neck cancer patients.^9^ This dataset was curated to support the development of the IMPACT (image-based mandibular and maxillary parcellation and annotation using computed tomography) tool.

This dataset provides a resource for evaluating dose-response relationships, testing auto-segmentation frameworks to support clinical decision-making, and developing predictive models of bone injury. It may also support interoperability efforts between dental and radiation oncology domains by providing a shared imaging reference standard.

## Methods

### Patient Cohort and Imaging Data

This dataset includes NIfTI (Neuroimaging Informatics Technology Initiative) files from diagnostic-quality simulation CT images, radiation dose distributions, and binary masks created from manually delineated sub-volumes of the mandible, maxilla, and individual teeth for 60 patients treated at The University of Texas MD Anderson Cancer Center for HNC. Patients were retrospectively selected from a philanthropically funded observational cohort (Stiefel Oropharynx Cancer Cohort, IRB: PA14-0947) under institutional review board approval (RCR030800). Inclusion criteria required availability of simulation CT, radiotherapy dose files, and dental structures of sufficient quality for manual contouring. Cases with extensive image artifacts or more than 20 missing teeth were excluded. Image data were exported from the RayStation (RaySearch Laboratories, Sweden) treatment planning system as standardized DICOM files and then converted to NIfTI and processed for its use in model development.

### Segmentation Protocol

Delineations were created on radiotherapy simulation CT scans using the RayStation treatment planning system. A schematic overview of the workflow is shown in Figure 1. The segmentation protocol was developed to account for anatomical and biological differences between basal and alveolar bone regions, incorporating laterality and tooth-specific labelling, offering compatibility with the recently proposed ASCO-endorsed ORN staging system, the ClinRad system.^10^Structures included 12 jaw sub-volumes (6 mandible, 6 maxilla) and up to 32 individual teeth per patient. A schematic overview of these sub-volumes is shown in Figure 2. Each jaw was divided into left, right, and central compartments, further separated into alveolar and basal regions. Sub-volume boundaries were defined using a consistent anatomical protocol, with alveolar regions extending from the alveolar crest 5 mm inferiorly (mandible) or 3 mm superiorly (maxilla). Basal regions were derived from contracted bone contours to preserve spatial distinction (see Figure 3). Teeth were contoured individually and labeled using the American Dental Association Universal Numbering System (1–32). Teeth were not segmented if they were absent or severely degraded on CT, but gaps were preserved in the numbering to maintain anatomical accuracy. To ensure generalizability of the model on edentulous or semi-edentulous patients, empty tooth sockets in cases with missing teeth were included in the alveolar region by manually adjusting the teeth expansion contour (see Figure 4).

**Figure 1.**
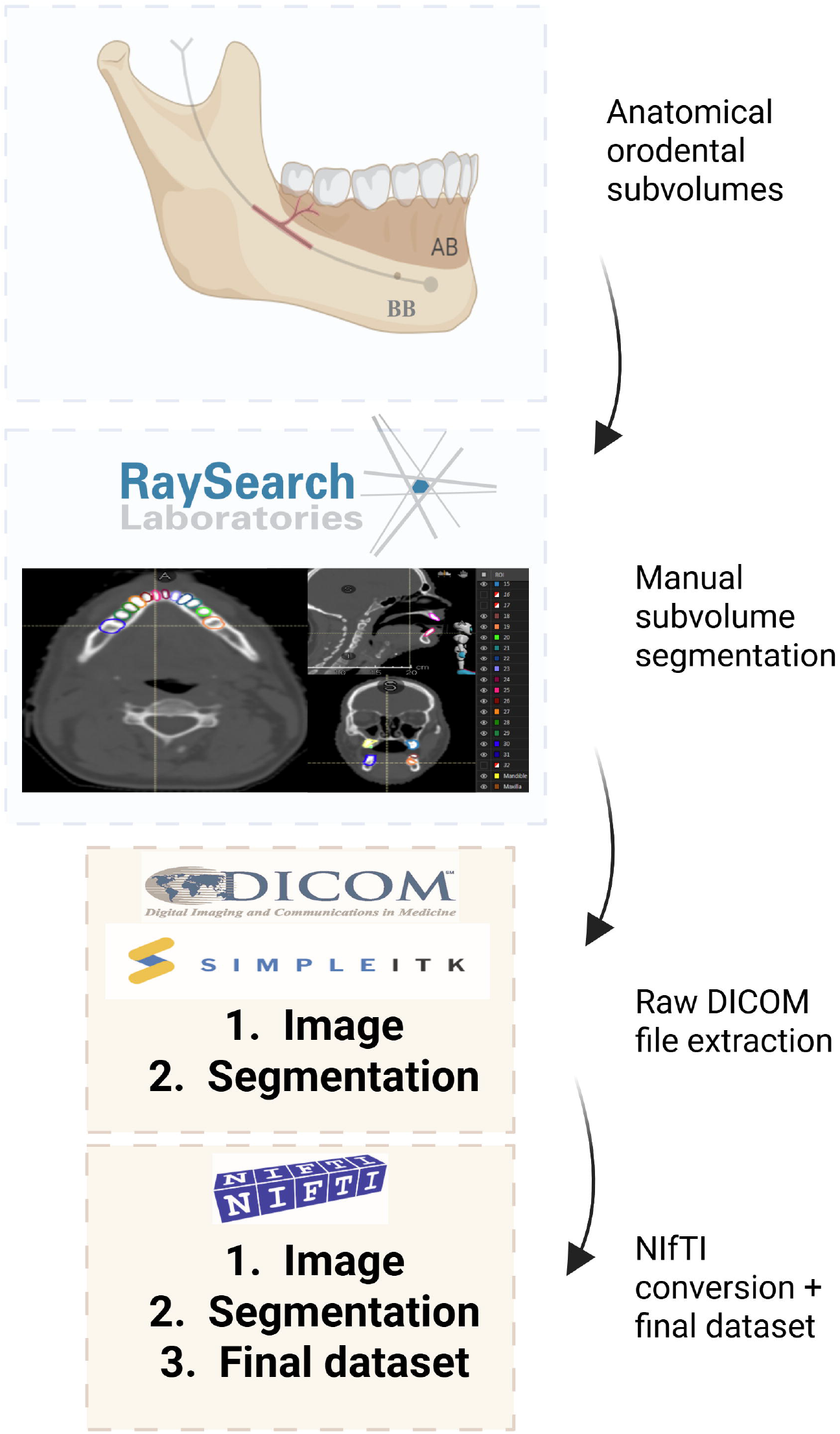
Data descriptor overview. Anatomical subvolumes were manually contoured in RayStation, followed by raw DICOM export. Imaging and segmentation data were converted to NIfTI format for downstream use.

**Figure 2.**
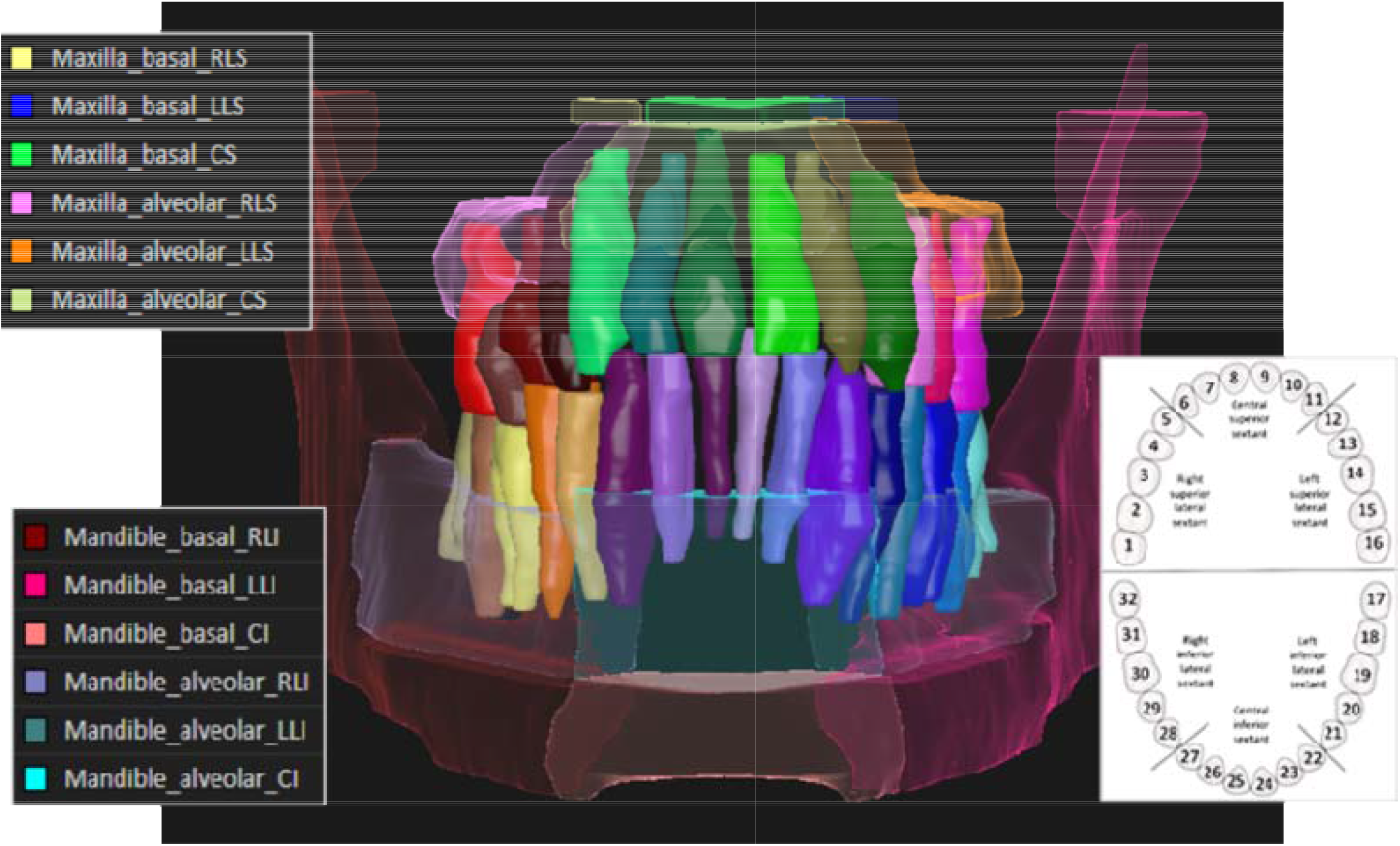
3D rendering of mandible and maxilla sub-volume segmentations. Alveolar and basal sub-volumes were manually contoured for the left, right, and central jaw, following anatomical landmarks and aligned with ClinRad ORN staging guidelines^10^. Color-coded labels indicate the structure, sub-region, and laterality.

**Figure 3.**
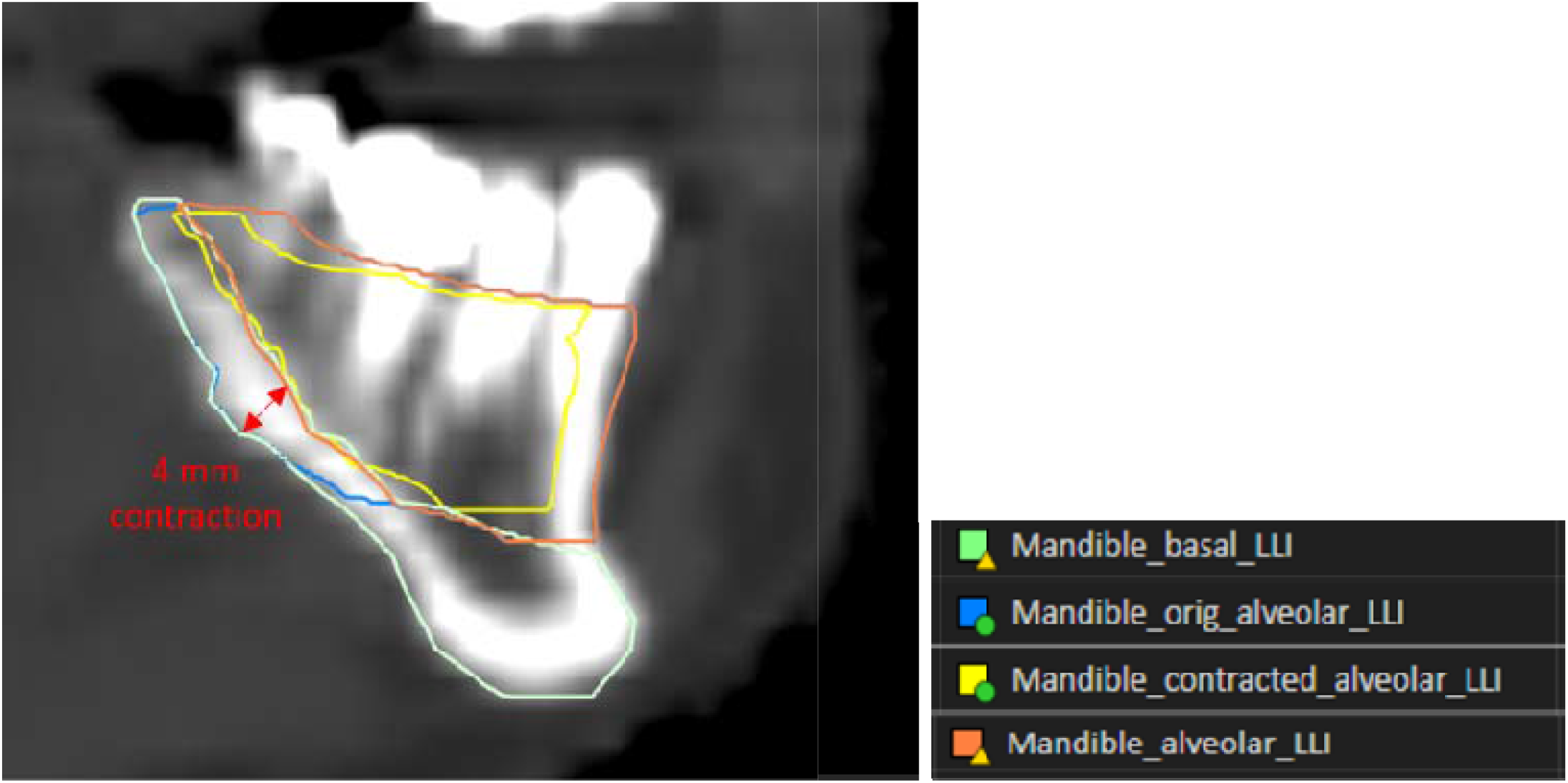
Contracted bone contours. Example demonstrating a 4 mm contraction of the mandible contour at the molars level to allow for a differentiation between basal and alveolar bone regions.

**Figure 4.**
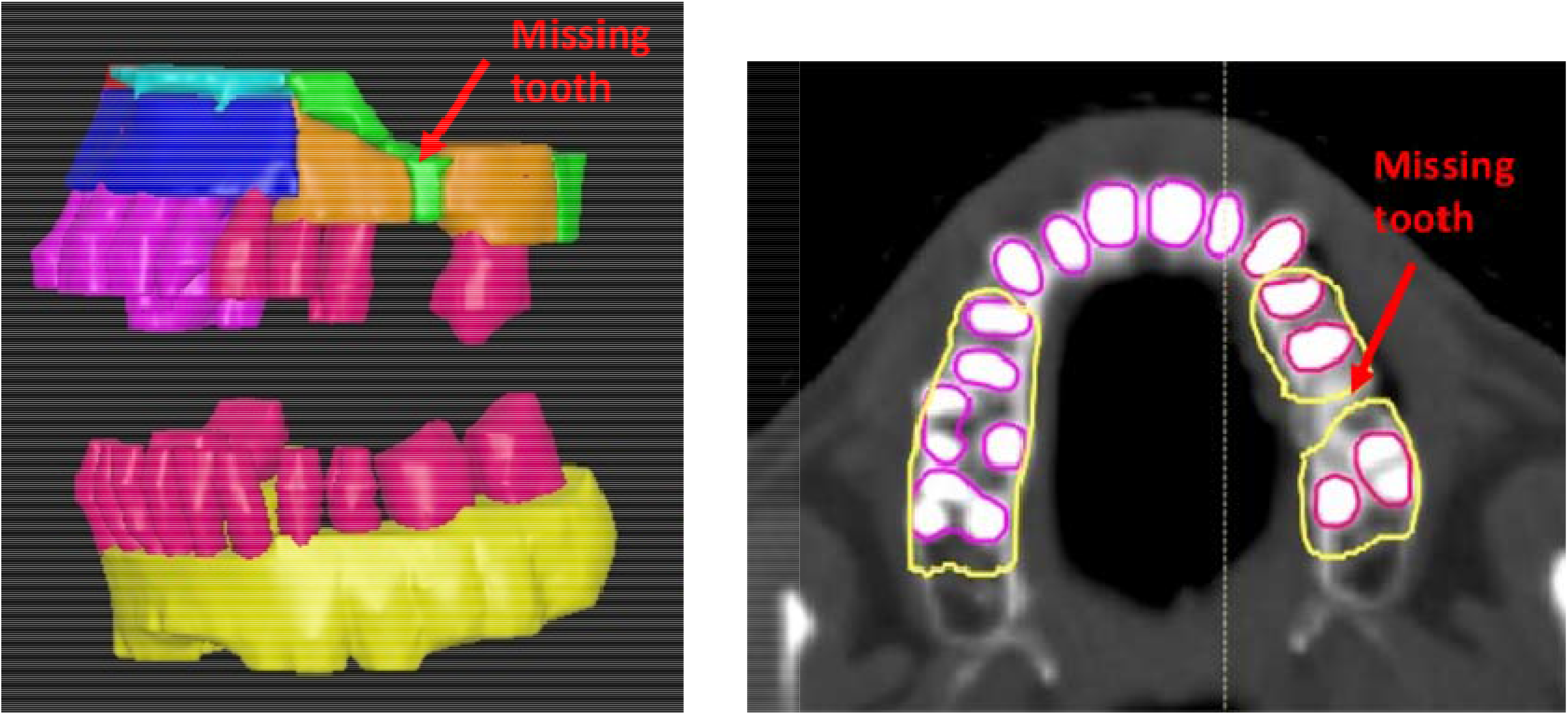
Missing teeth. Example of a case with a missing tooth. To ensure generalizability of the model on edentulous or semi-edentulous patients, empty tooth sockets in cases with missing teeth were included in the alveolar region by manually adjusting the teeth expansion contour.

### Data Conversion and Preprocessing

DICOM images and RT structure sets were exported and converted to compressed NIfTI format (.nii.gz), which preserves 3D spatial resolution and orientation, using SimpleITK (version: 2.2.1) and dcmrtstruct2nii (version: 2). Contours were rasterized into binary masks using a label-priority strategy (with central volumes having priority over lateral ones), ensuring consistent voxel assignments where overlaps occurred. For a subset of 8 test cases, radiation dose distribution DICOM files (converted to NIfTI) were available and included in this dataset. While further preprocessing steps were followed to prepare the data for model development,^9^ the shared dataset contains the unprocessed NIfTI files.

### Structure Naming Convention

Each segmented region follows a standardized naming schema based on three components: anatomical structure (mandible or maxilla), bone type (alveolar or basal), and location (left, right, central). For example, Mandible_alveolar_RLI refers to the right lateral inferior alveolar region of the mandible. Teeth masks were labeled with numbers (1, 2, etc.) based on ADA numbering.

### Data Records

All data records are publicly available through Figshare at https://doi.org/10.6084/m9.figshare.28615874 are organized by patient ID in a standardized folder hierarchy (see Figure 5). Each patient folder contains planning CT images, dose distributions, and structure masks in NIfTI format.

**Figure 5.**
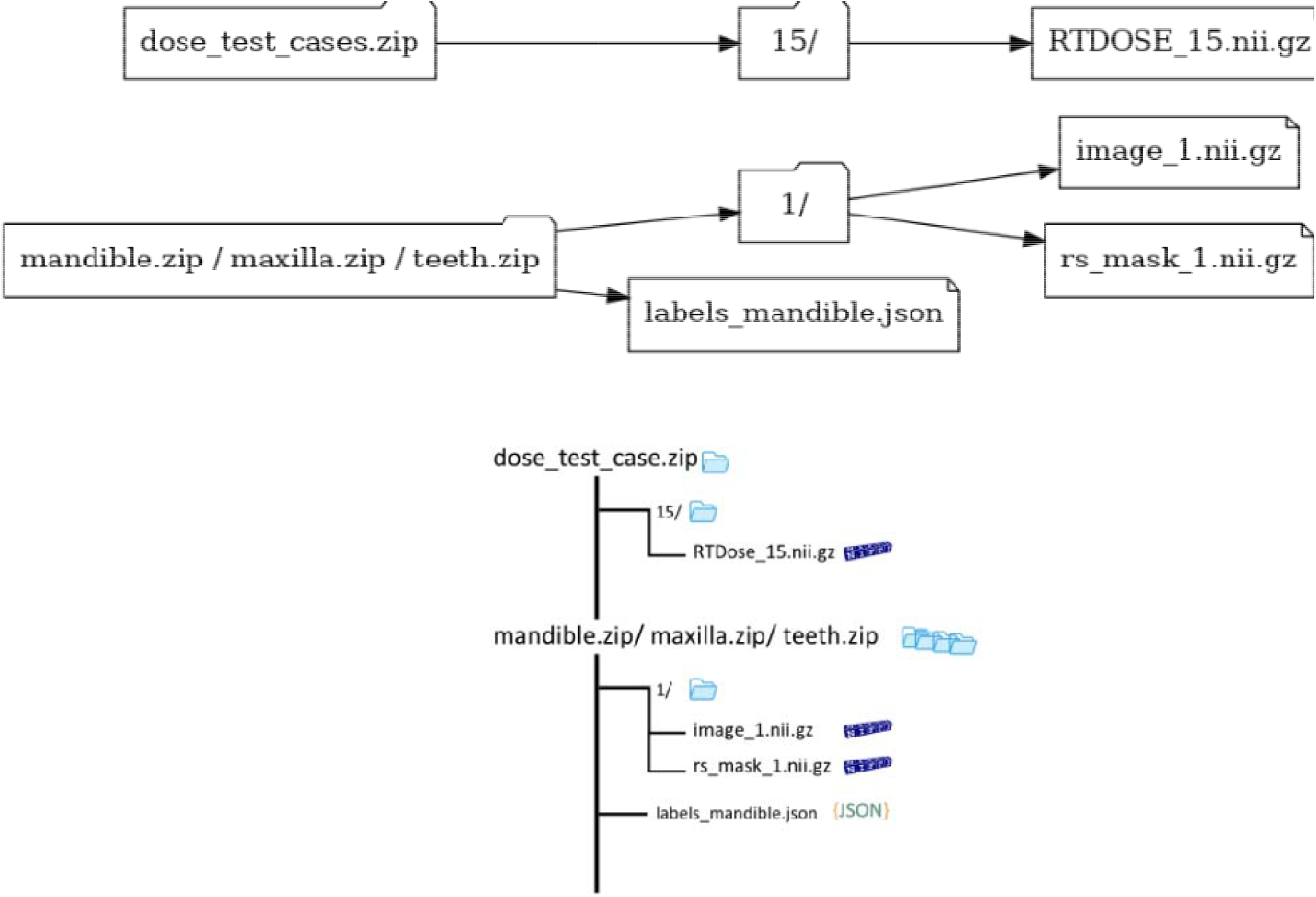
Directory structure of the IMPACT_ORN dataset. The dataset is organized into four archives: mandible.zip, maxilla.zip, teeth.zip, and dose_test_cases.zip. Each anatomical archive contains patient-specific folders with a planning CT image (image_<ID>.nii.gz) and corresponding segmentation mask (rs_mask_<ID>.nii.gz). A labels_<structure>.json file at the root of each archive provides label definitions. The dose archive contains a subset of RT dose files (RTDOSE_<ID>.nii.gz) organized by patient ID for a subset of test patients. All files are provided in standardized .nii.gz (NIfTI) or .json formats.

Each anatomical dataset archive (mandible.zip, maxilla.zip, and teeth.zip) contains a collection of patient folders, each named by patient ID (e.g., 1/, 10/). Within each folder are two primary files: a planning CT scan (image_<ID>.nii.gz) and a corresponding segmentation mask (rs_mask_<ID>.nii.gz). A separate archive (dose_test_cases.zip) contains RT dose distributions for a subset of patients, structured similarly, with each folder containing a dose file (RTDOSE_<ID>.nii.gz). At the root level of each anatomical archive, a JSON file (e.g., labels_mandible.json) defines the label names and indices used in the segmentation masks. All files are stored in compressed NIfTI (.nii.gz) or JSON format and follow a consistent naming convention to support reproducibility and automated processing. A visual overview of the dataset directory structure is provided in Figure 5.

### Technical Validation

Structures were manually contoured by 10 observers of varying degree of expertise, including undergraduate (SJ, PG) and postgraduate (SM, JC, SA, AS, PO) students and doctoral (NW, ZK) and postdoctoral (AMSA) trainees. Observers followed detailed contouring instructions and an associated structure set template to achieve consistency in the contouring methodology previously agreed amongst a group of clinical experts (LHV, ACM, CDF, EW, AH, RAW). Finally, all contours were reviewed by two clinical experts (LHV, ACM).

Automated quality control checks were applied to verify contour completeness, laterality labeling, and consistent spatial alignment between structure masks and the CT volume. Overlapping regions were resolved using a defined label-priority system to ensure that each voxel was assigned to a single anatomical class. All exported masks were visually inspected in 3D Slicer to confirm structural fidelity and anatomical plausibility.

## Data Availability

All data records are publicly available through Figshare at https://doi.org/10.6084/m9.figshare.28615874 are organized by patient ID in a standardized folder hierarchy (see Figure 5).
Custom code used for image preprocessing related to the dataset described is available at https://github.com/Acasteo/IMPACT_dcm_to_nii. DICOM images and RT structure sets were exported and converted to compressed NIfTI format (.nii.gz), which preserves 3D spatial resolution and orientation, using SimpleITK and dcmrtstruct2nii. Contours were rasterized into binary masks using a label-priority strategy (with central volumes having priority over lateral ones), ensuring consistent voxel assignments where overlaps occurred. The repository includes scripts for converting DICOM RTSTRUCTs to NIfTI format and creating the binary masks. All code was developed and tested using Python 3.9

https://github.com/Acasteo/IMPACT_dcm_to_nii

## Code Availability

Custom code used for image preprocessing related to the dataset described is available at https://github.com/Acasteo/IMPACT_dcm_to_nii. DICOM images and RT structure sets were exported and converted to compressed NIfTI format (.nii.gz), which preserves 3D spatial resolution and orientation, using SimpleITK and dcmrtstruct2nii. Contours were rasterized into binary masks using a label-priority strategy (with central volumes having priority over lateral ones), ensuring consistent voxel assignments where overlaps occurred. The repository includes scripts for converting DICOM RTSTRUCTs to NIfTI format and creating the binary masks. All code was developed and tested using Python 3.9

## Acknowledgements

This work was supported directly or in part by personnel funding/resource support from the National Institutes of Health (NIH) National Institute for Dental and Craniofacial Research (K01DE030524, U01DE032168, R21DE031082, R56/R01DE025248, R01DE028290); NIH National Cancer Institute (P01CA285249, K12CA088084, P30CA016672); the University of Texas MD Anderson Cancer Center Charles and Daneen Stiefel Center for Head and Neck Cancer Oropharyngeal Cancer Research Program; and the MD Anderson Image-guided Cancer Therapy Program. Additionally, M.A.N. received funding from the National Institutes of Health/National Institute of Dental and Craniofacial Research (NIH/NIDCR) through grant 1R03DE033550-01. C.D.F. and S.Y.L. received related funding support from the NIH/NIDCR (U01DE032168/ R01DE025248). C.D.F. also receives infrastructure and salary support through the NIH/NCI MD Anderson Cancer Center Core Support Grant (CCSG) Image-Driven Biologically-informed Therapy (IDBT) program (P30CA016672-47). S.Y.L. is supported through the CCSG Head and Neck Program (P30CA016672-48). K.A.W. was supported by an Image Guided Cancer Therapy (IGCT) T32 Training Program Fellowship from T32CA261856. L.V.V. received funding and salary support from KWF Dutch Cancer Society through a Young Investigator Grant (KWF-13529) and from NWO ZonMw through the VENI grant (NWO-09150162010173). K.K.B. and A.H.C. acknowledge support from the Image Guided Cancer Therapy Research Program at The University of Texas MD Anderson Cancer Center, which was partially funded by the National Institutes of Health/NCI under award number P30CA016672 and through a generous gift from the Apache Corporation. S.J.F. received funding from Hitachi, NIH/NCI, the National Association of Proton Therapy, Affirmed Pharma and NASA/Baylor College of Medicine and honoraria from IBA. N.A.W. was supported by a training fellowship from UTHealth Houston Center for Clinical and Translational Sciences T32 Program (Grant No. T32 TR004905) and a NIH National Institute of Dental and Craniofacial Research (NIDCR) Academic Industrial Partnership Grant (R01DE028290), and the American Legion Auxiliary Fellowship in Cancer Research. S.J. acknowledges funding support from 1 R25 CA 265800-1 A1. Z.K. was supported by a doctoral fellowship from the Cancer Prevention Research Institute of Texas grant RP210042. Parshan Gerafian or P.G was supported by the 1R25CA240137 UPWARDS Training Program and P.G. was also supported by the CPRIT Research Training Award CPRIT Training Program (RP210028).

## Author contributions

Conceptualization: LHV, AHC, RH, CDF, MAN, ACM. Methodology: LHV, AHC, RH, LVV, KAW, AH, EW, RAW, CDF, MAN, ACM. Software: LHV, AHC, RH, CDF, MAN, ACM. Validation: LHV, AHC, RH, CDF, MAN, ACM. Formal analysis: LHV, AHC, RH, CDF, MAN, ACM. Investigation: LHV, AHC, RH, DJR, CW, HCW, CDF, MAN, ACM. Resources: DJR, HCW, MC, KKB, SYL, CDF, MAN, ACM. Data curation: LHV, AHC, RH, DJR, SJ, PG, NAW, ZK, SM, JC, SA, AS, PO, AMSA, MAN, ACM. Writing - original draft: LHV, AHC, RH, CDF, MAN, ACM. Writing - review and editing: LHV, AHC, RH, LVV, DJR, CW, HCW, KAW, SJ, PG, NAW, ZK, SM, JC, SA, AS, PO, AMSA, AH, EW, RAW, SJF, CEAB, KKB, MSC, MW, KAH, SYL, CDF, MAN, ACM. Visualization: LHV. Supervision: CDF, ACM. Project administration: ACM. Funding: CDF, KKB, MAN, ACM.

## References

(1) Qiu, B.; Der Wel, H. V.; Kraeima, J.; Glas, H. H.; Guo, J.; Borra, R. J. H.; Hendrikus Witjes, M. J.; Van Ooijen, P. M. A. Automatic Segmentation of Mandible from Conventional Methods to Deep Learning-a Review. Journal of Personalized Medicine 2021, 11 (7). 10.3390/jpm11070629.

(2) Kargilis, D. C.; Xu, W.; Reddy, S.; Ramesh, S. S. K.; Wang, S.; Le, A. D.; Rajapakse, C. S. Deep Learning Segmentation of Mandible with Lower Dentition from Cone Beam CT. Oral Radiol 2025, 41 (1), 1–9. 10.1007/s11282-024-00770-6.

(3) Verhelst, P.-J.; Smolders, A.; Beznik, T.; Meewis, J.; Vandemeulebroucke, A.; Shaheen, E.; Van Gerven, A.; Willems, H.; Politis, C.; Jacobs, R. Layered Deep Learning for Automatic Mandibular Segmentation in Cone-Beam Computed Tomography. Journal of Dentistry 2021, 114, 103786. 10.1016/j.jdent.2021.103786.

(4) Cui, Z.; Fang, Y.; Mei, L.; Zhang, B.; Yu, B.; Liu, J.; Jiang, C.; Sun, Y.; Ma, L.; Huang, J.; Liu, Y.; Zhao, Y.; Lian, C.; Ding, Z.; Zhu, M.; Shen, D. A Fully Automatic AI System for Tooth and Alveolar Bone Segmentation from Cone-Beam CT Images. Nat Commun 2022, 13 (1), 2096. 10.1038/s41467-022-29637-2.

(5) Tao, S.; Wang, Z. Tooth CT Image Segmentation Method Based on the U-Net Network and Attention Module. Computational and Mathematical Methods in Medicine 2022, 2022 (1), 3289663. 10.1155/2022/3289663.

(6) Cui, Z.; Li, C.; Wang, W. ToothNet: Automatic Tooth Instance Segmentation and Identification From Cone Beam CT Images. 2019 IEEE/CVF Conference on Computer Vision and Pattern Recognition (CVPR) 2019, 6361–6370. 10.1109/CVPR.2019.00653.

(7) Gao, H.; Chae, O. Individual Tooth Segmentation from CT Images Using Level Set Method with Shape and Intensity Prior. Pattern Recognition 2010, 43 (7), 2406–2417. 10.1016/j.patcog.2010.01.010.

(8) Jang, T. J.; Kim, K. C.; Cho, H. C.; Seo, J. K. A Fully Automated Method for 3D Individual Tooth Identification and Segmentation in Dental CBCT. IEEE Trans Pattern Anal Mach Intell 2022, 44 (10), 6562–6568. 10.1109/TPAMI.2021.3086072.

(9) Humbert-Vidan, L.; Castelo, A. H.; He, R.; van Dijk, L. V.; Rhee, D. J.; Wang, C.; Wang, H. C.; Wahid, K. A.; Joshi, S.; Gerafian, P.; West, N.; Kaffey, Z.; Mirbahaeddin, S.; Curiel, J.; Acharya, S.; Shekha, A.; Oderinde, P.; Ali, A. M. S.; Hope, A.; Watson, E.; Wesson-Aponte, R.; Frank, S. J.; Barbon, C. E. A.; Brock, K. K.; Chambers, M. S.; Walji, M.; Hutcheson, K. A.; Lai, S. Y.; Fuller, C. D.; Naser, M. A.; Moreno, A. C. Image-Based Mandibular and Maxillary Parcellation and Annotation Using Computer Tomography (IMPACT): A Deep Learning-Based Clinical Tool for Orodental Dose Estimation and Osteoradionecrosis Assessment. 2025. 10.1101/2025.03.18.25324199.

(10) Watson, E. E.; Hueniken, K.; Lee, J.; Huang, S. H.; El Maghrabi, A.; Xu, W.; Moreno, A. C.; Tsai, C. J.; Hahn, E.; McPartlin, A. J.; Yao, C. M. K. L.; Goldstein, D. P.; De Almeida, J. R.; Waldon, J. N.; Fuller, C. D.; Hope, A. J.; Ruggiero, S. L.; Glogauer, M.; Hosni, A. A. Development and Standardization of an Osteoradionecrosis Classification System in Head and Neck Cancer: Implementation of a Risk-Based Model. J Clin Oncol 2024, 42 (16), 1922–1933. 10.1200/JCO.23.01951.

